# Blood Cytokine Profiling Reveals Distinct Inflammatory Signatures in Phenotypes A and B of Polyendocrine Metabolic Ovarian Syndrome (PMOS; formerly PCOS)

**DOI:** 10.64898/2026.10.20.26365978

**Authors:** Lipika Priyadarsini Patra, Betcy Susan Johnson, K Jayakrishnan, Sathy M Pillai, Malini Laloraya

**Affiliations:** Female Reproduction and Metabolic Syndromes Laboratory, Division of Molecular Reproduction, Rajiv Gandhi Centre for Biotechnology, Thiruvananthapuram −695 014, Kerala, India; Regional Centre for Biotechnology, Pali, Faridabad-121001, Haryana, India; KJK Hospital, Mar Ivanios College Road, Nalanchira, Thiruvananthapuram, 695015, Kerala, India; SAMAD IVF Hospitals, V. V. Road, Pattoor, Thiruvananthapuram −695 035, Kerala, India

**Keywords:** Polyendocrine metabolic ovarian syndrome (PMOS), Cytokine profiling, Immune dysregulation, Inflammatory biomarkers, Phenotypic heterogeneity

## Abstract

Polyendocrine Metabolic Ovarian Syndrome (PMOS, formerly PCOS) is associated with chronic low-grade inflammation. This study aimed to characterize the peripheral cytokine landscape in women with PMOS and identify cytokines associated with disease pathogenesis and phenotypic variation. Plasma concentrations of 48 cytokines were quantified using a multiplex bead-based assay in 102 women with PMOS and 114 healthy controls. We performed receiver operating characteristic (ROC) analysis, odds ratio estimation, correlation analysis, and network analyses to characterise cytokine alterations and identify phenotype-associated inflammatory signatures. Fourteen cytokines were significantly elevated in PMOS compared with healthy controls: IL-18, IL-1Ra, IL-12p70, IL-13, IL-16, MCP-1, MCP-3, CTACK, Eotaxin, RANTES, IP-10, M-CSF, HGF, and β-NGF. ROC analysis identified IL-16, CTACK, and IP-10 as potential discriminators of phenotype A, whereas IL-18, M-CSF, and HGF demonstrated promising discriminatory performance for phenotype B. Odds ratio analysis identified 11 cytokines with odds ratios greater than 1, with IL-16 showing the strongest association with PMOS. Correlation analysis revealed 10 strongly correlated cytokine pairs (r ≥ 0.7) in the PMOS group, suggesting coordinated regulation of inflammatory mediators. Gene ontology and network analysis highlighted alterations in chemotaxis and immune cell migration pathways, implicating these processes in PMOS-associated immune dysregulation. These findings reveal a distinct peripheral inflammatory cytokine signature and phenotype-associated differences in PMOS. IL-16 emerged as a prominent candidate for further investigation, given its strong association with PMOS. Peripheral cytokine profiling may provide a minimally invasive approach to characterising immune alterations and identifying candidate biomarkers for disease characterisation and phenotype stratification.

Graphical Abstract:
Peripheral Cytokine Signatures Reveal Immune Dysregulation and Phenotypic Heterogeneity in PMOS

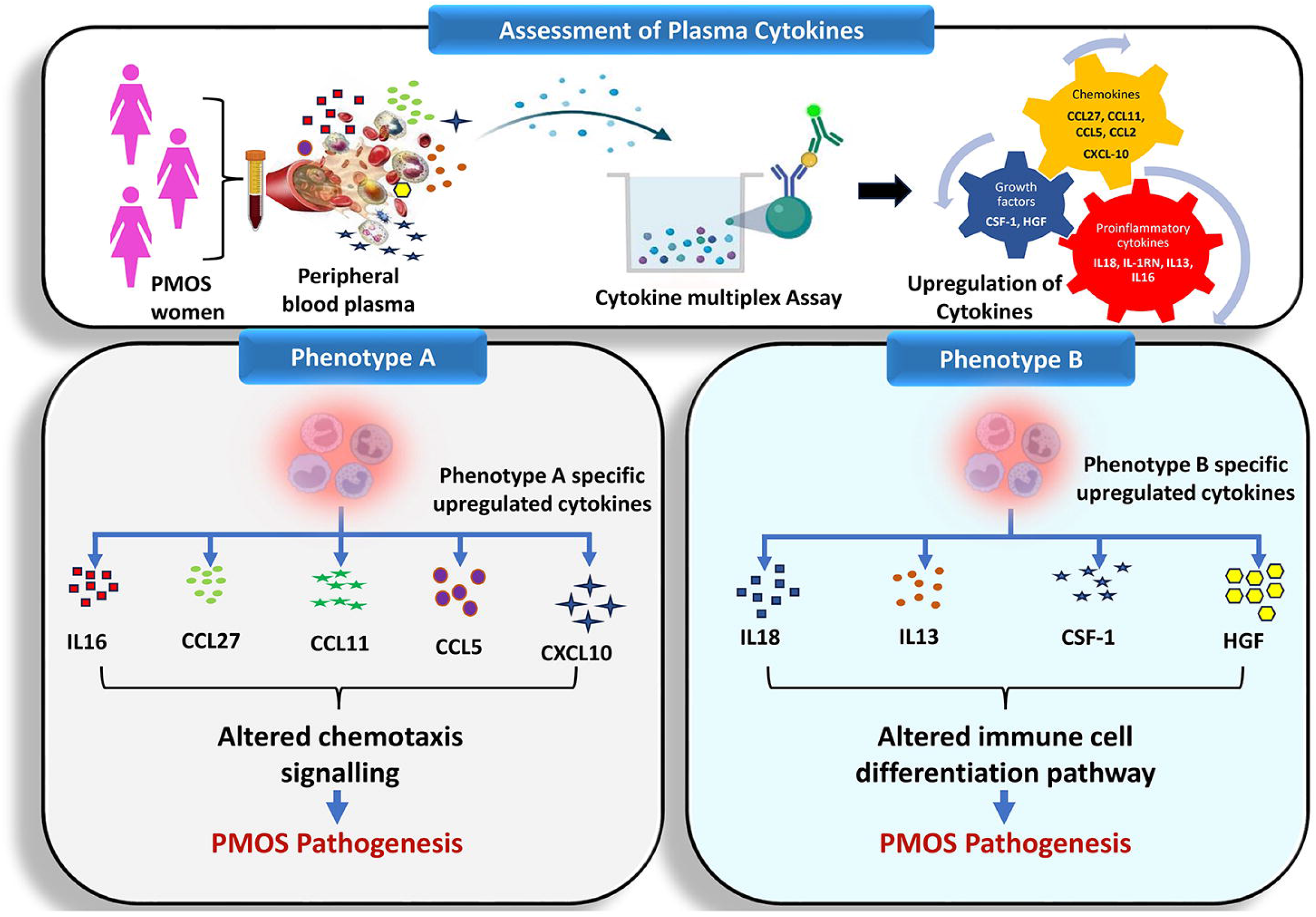

## INTRODUCTION

Polycystic Metabolic Ovarian Syndrome (PMOS), formerly known as Polycystic Ovary Syndrome (PCOS), is a familiar endocrine disorder that affecting women of reproductive age, with a prevalence of 15%-20%, depending on various diagnostic criteria (1, 2). It is characterized by hyperandrogenaemia, polycystic ovaries, and oligo/anovulation (3) and is frequently associated with insulin resistance, Type-2 diabetes, visceral obesity, hirsutism, and acne. Increasing evidence suggests that chronic low-grade inflammation contributes significantly to PMOS pathogenesis. Elevated levels of inflammatory markers, including C-reactive protein (CRP) (4), CRP/albumin ratio (5) and increased leukocyte counts (6) have been consistently reported in PMOS patients, independent of obesity, highlighting inflammation as a key feature of the disorder. Furthermore, inflammatory processess are thought to link hyperandrogenism and insulin resistance with the metabolic and cardiovascular complications of PMOS (7).

A pro-inflammatory environment causes the release of cytokines which are critical mediators of immune communication and play vital role in reproductive physiology including steroidogenesis (8), folliculogenesis, ovulation (9), and endometrial remodeling (10). Cytokine production is tightly regulated by sex steroids, resulting in a gender-specific immune response (11). Emerging evidence indicates that immune dysregulation contributes to PMOS development; however, studies investigating cytokines in PMOS have largely focused on a limited number of inflammatory mediators, often yielding contradictory results (12). Consequently, a comprehensive characterization of the PMOS cytokine landscape remains lacking.

Because circulating cytokines can be measured non-invasively and may serve as clinically useful biomarkers, systematic profiling of cytokine networks may provide valuable insights into the immune mechanisms underlying PMOS. Therefore, in the present study, we performed comprehensive plasma cytokine profiling in women with PMOS and healthy controls using the Bio-Plex Pro™ Human Cytokine Screening 48-Plex Panel. This approach enabled the simultaneous assessment of interleukins, chemokines, and growth factors to identify immune signatures associated with PMOS pathogenesis and phenotypic heterogeneity.

## 1. Materials and Methods

### 1.1. Subject recruitment

This retrospective longitudinal study included 102 women with Polyendocrine Metabolic Ovarian Syndrome (PMOS) and 114 age-matched healthy controls (18–40 years) recruited from the extreme southern region of India (Kerala). PMOS was diagnosed according to the Rotterdam ESHRE/ASRM consensus criteria. Healthy controls had regular menstrual cycles, no history of metabolic or hormonal disorders, and no fertility-related complications. Individuals with other androgen disorders, autoimmune diseases, or recent infections (<3 weeks) were excluded.

**Ethics statement:** The Institutional Ethics Committee of the Rajiv Gandhi Centre for Biotechnology (RGCB), Thiruvananthapuram, gave ethical approval for the study (Approval Nos. IHEC/01/2019/07 and IHEC/01/2015/09). Written informed consent was obtained from all participants in compliance with the Declaration of Helsinki.

### 1.2. Sample collection and processing

Peripheral blood (5 mL) was collected in EDTA-coated tubes (CML Biotech (P) Ltd., Code VS-K3) during days 2–5 of the follicular phase. Plasma was isolated following 3% dextran-mediated erythrocyte sedimentation and centrifugation during PBMC preparation. Samples were aliquoted and stored at −80°C until analysis. Each aliquot was thawed only once before use.

### 1.3. Hormone profiling by ELISA

Plasma concentrations of luteinizing hormone(LH), follicle stimulating hormone(FSH), testosterone and Dihydrotestosterone(DHT) were estimated using commercially available ELISA kits according to the manufacturers’ instructions. (LH-#500720, FSH-#500710, Testosterone-#582701 from Cayman Chemical, MI, USA and DH-#MBS2000056, MyBiosource Inc, CA, USA).

### 1.4. Cytokine profiling by Bioplex assay

Plasma levels of 48 cytokines were quantified using the Bio-Plex Pro^Tm^ Human Cytokine Screening 48 plex panel (Cat #12007283, Bio-Rad Laboratories, CA, USA). Assays were performed according to the manufacturer’s protocoland analyzed using a BioPlex 200 instrument equipped with Bio-Plex Manager analysis software Version 6 (Bio-Rad Laboratories, Hercules, CA, USA). BioPlex 200 instrument setting for cytokine analysis was: 50 beads per region, 60μl/min flow rate, and 5000 - 25000 discriminator gate. Cytokine concentrations were calculated from standard generated using recombinant cytokine standards included in the kit. Identical positive quality controls were also included in the assay. The values with <OOR were taken as zero, values with >OOR were taken as the highest concentration values present inside the standard graph of that particular cytokine, while values with * are machine extrapolated values, and thus, included in our study group.

### 1.5. Statistical Analysis

Cytokine concentrations(pg/ml) were calculated using a five-parameter logistic regression model. Statistical analyses were performed using GraphPad Prism 8. Differences between PMOS and control groups were assessed using Student’s *t*-test, while phenotype-specific comparisons were performed using one-way ANOVA followed by Tukey’s multiple-comparison test. Receiver operating characteristic (ROC) analysis was conducted using MetaboAnalyst 6.0 to evaluate the diagnostic potential of altered cytokines. Odds ratios (ORs) with 95% confidence intervals were calculated and visualized using forest plots. Further, Pearson’s correlation analysis was performed for cytokines with OR>1, and correlation heat maps were generated. Linear regression analysis was used to assess associations between cytokine levels and BMI. Statistical significance was defined as *p* < 0.05.

### 1.6. Gene Ontology and network analysis

Cytokines exhibiting an OR>1 were subjected to Gene Ontology (GO) and network enrichment analysis using the GOnet platform. Log2-transformed cytokine expression values were used to identify cytokine–cytokine interactions, biological processes, and signaling pathways associated with PMOS. Poorly connected nodes were excluded, and significantly enriched networks were retained for downstream analysis.

## 2. Results

All subjects included in our study are of similar age groups. The body mass index (BMI) of the PMOS group is significantly high compared to the control. The clinical and physiological characteristics of subjects enrolled in our study were reported in Table 1. No change in FSH level between the two groups is seen. The level of LH is significantly high in PMOS patients (p-value <0.0001). LH/FSH ratio is significantly high in PMOS patients compared to controls (p-value <0.0001). The total testosterone level (p-value <0.0001) as well as DHT levels (p-value <0.01) are significantly increased in PMOS women. The flow diagram detailing the step-by-step method followed is presented in Figure 1.

**Figure 1:**
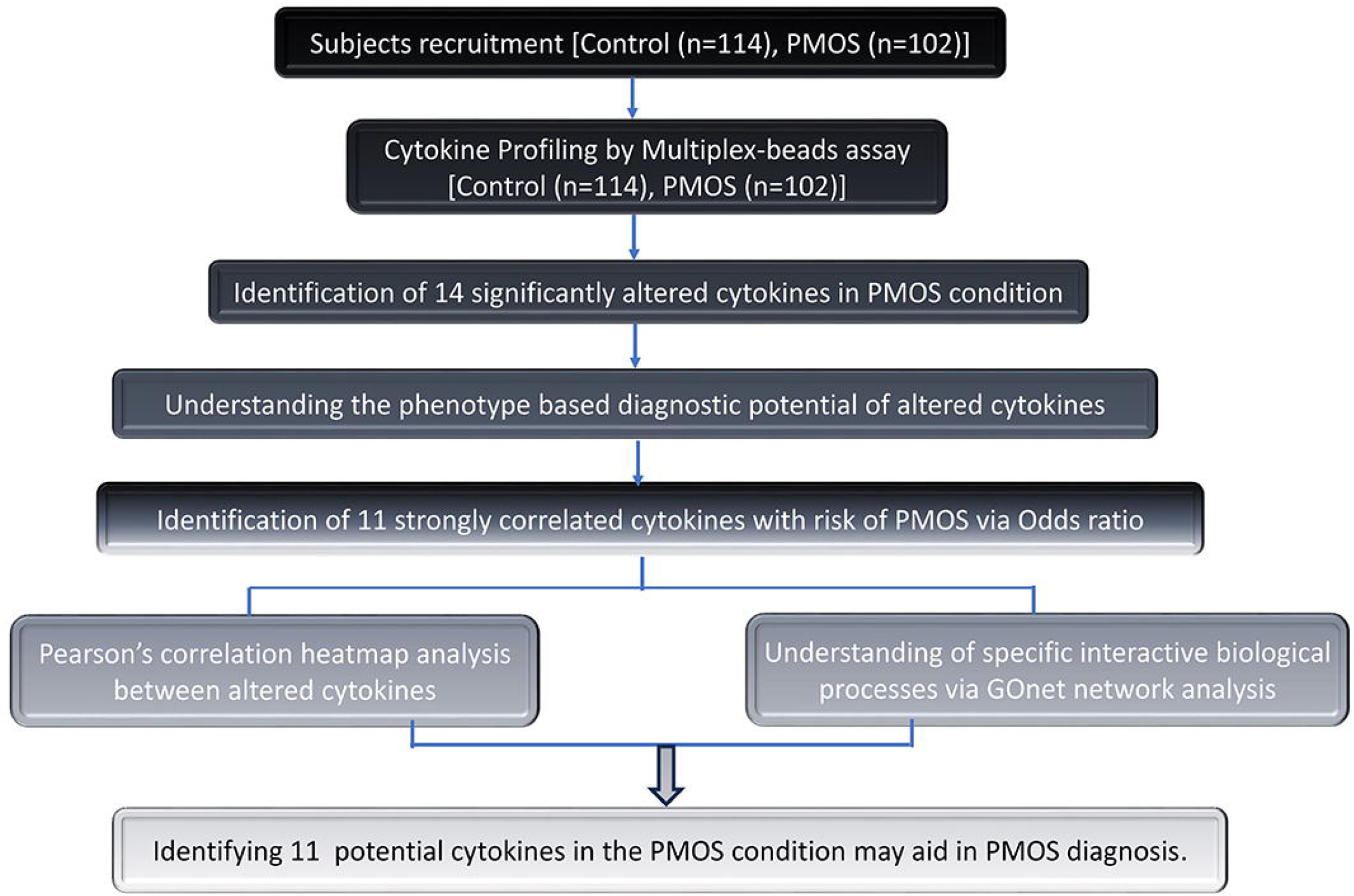
Flow chart of the methodology used for the work. Flowchart showing the recruitment of total control and PMOS subjects and screening of 11 potential cytokines in PMOS condition via various statistical analyses.

**Table 1.** Clinical and physiological characteristics features of Control (n=114) and PCOS (n=102) subjects.

| Characteristics features | Control (n=114) | PCOS (n=102) | P value |
| --- | --- | --- | --- |
| Age (years) | 26.8±0.35 | 27.6±0.36 | P value = 0.129 |
| BMI (kg/m<sup>2</sup>) | 22.6±0.32 | 25.6±0.4 | P value <0.0001 |
| LH (mIU/ml) | 7.1± 0.94 | 15.1 ± 0.6 | P value< 0.0001 |
| FSH (mIU/ml) | 6.6± 0.27 | 6 ± 0.34 | P value = 0.233 |
| LH/FSH | 1.1± 0.17 | 3 ± 0.17 | P value < 0.001 |
| Total Testosterone (ng/dl) | 33.4±1.79 | 46.3 ± 1.74 | P value <0.0001 |
| DHT (ng/mL) | 0.1± 0.01 | 0.2 ± 0.01 | P value <0.01 |
| Menstrual cycle length | 28-35 days | Menstrual cycle > 40 days |  |

**Footnote for Table 1:** All values were presented as mean± SEM.

### 2.1. Phenotypic classification of PMOS subjects included in the study group

The phenotypic classification of PMOS was introduced by the Androgen excess-PMOS phenotype Task Force based on NIH/NICHD Rotterdam criteria in 1990 and 2003 (13). PMOS individuals (n=102) included in this study were characterized into four main phenotypic subgroups: phenotype A, B, C, and D based on the diagnostic criteria of hyperandrogenism (HA), Ovulatory Dysfunction (OD) and Polycystic Ovarian Morphology (PCOM). Our study group showed that phenotype A (HA+OD+PCOM) is the most prevalent subgroup with 52.9% of cases. While phenotype B (HA+OD) is the least found subgroup with 4.9% cases. We found that phenotype C (HA+PCOM) is the second highest phenotype with 31.4% cases and phenotype D (OD+PCOM) with 10.8% cases in the extreme southern part of India (Kerala). There is no statistically significant difference in the levels of LH, FSH, LH/FSH, and DHT among the four PMOS phenotypic populations of our study group. We observed a significantly higher level of testosterone in the population of phenotype A, B, and C compared to phenotype D. Reproductive complications were observed in all four groups. The clinical and physiological characteristics in each PMOS phenotype is given in the Supplementary Table S2.

### 2.2. Plasma cytokines show differential expression in PMOS patients

Out of 48 cytokines measured, nine cytokines showed statistically significant differential expression as depicted in (Figure 2A-N). Levels of 14 cytokines that showed variation were listed in Table 2. The mean level of interleukin 1 family members IL18 (p-value<0.01) and IL-1Ra (p-value <0.001) were significantly increased in PMOS patients compared to controls (Figures 2A-2B). The level of Th1 cytokine-like IL12p70 is significantly high in PMOS conditions (p-value< 0.05) (Figure 2C). Also, Th2 cytokine-like IL13 (p-value<0.05) is significantly elevated in PMOS conditions (Figure 2D). Proinflammatory cytokines IL16 (p-value<0.01) are also significantly high in PMOS conditions (Figure 2E). Plasma chemokines like MCP-1(CCL2) (p-value<0.0001), MCP-3 (CCL7) (p-value<0.05), CTACK (CCL27) (p-value <0.01), Eotaxin (CCL11) (p-value<0.05) and RANTES (CCL5) (p-value<0.01) (Figure 2F-2J) were significantly increased in PMOS patients. Additionally, CXC-motif chemokine like IP-10 (CXCL-10) (p-value<0.05), was also increased significantly in PMOS women compared to control (Figure 2K). PMOS groups showed a significantly higher level of macrophage-colony stimulating factors like M-CSF(CSF-1) (p-value<0.01), growth factors like HGF (p-value<0.05) and β-NGF (p-value<0.05) were also elevated in PMOS women compared to control (Figure 2L-2N). The plasma concentration of the other thirty-four cytokines didn’t reveal a significant difference between PMOS and control groups as shown in Supplementary Table S1 and Supplementary Figure S1.

**Figure 2:**
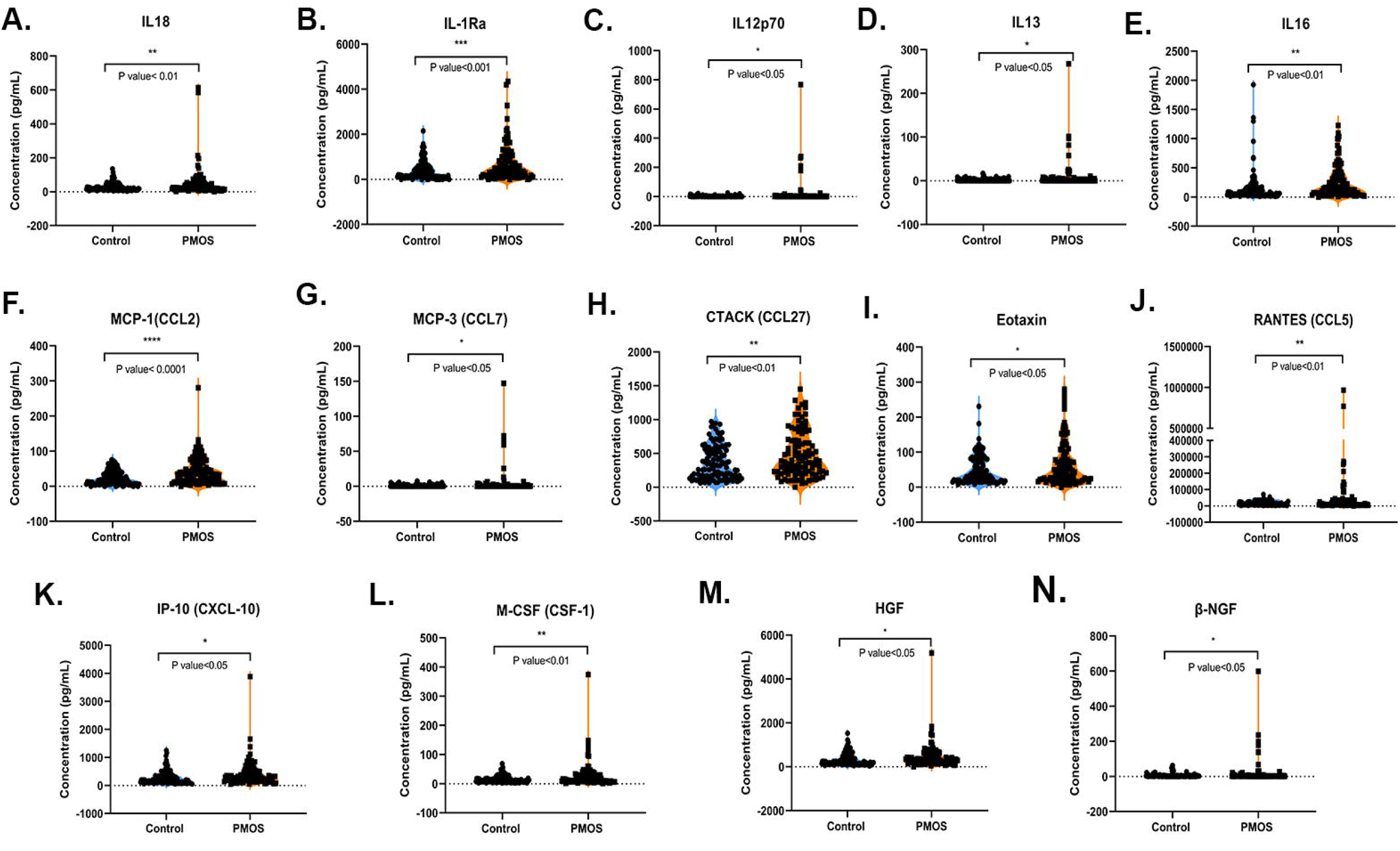
Comparative analysis of various significantly altered cytokines in control and PMOS groups. Comparison of mean cytokine concentration (pg/mL) change in control and PMOS groups. (A-N) shows scatter dot plot representation of the significantly altered cytokines in control and PMOS groups. Round dots represent control subjects and square dots represent PMOS subjects. Each dot is represented as a single subject and a p-value < 0.05 is considered statistically significant.

**Table 2.** Levels of differential cytokines in Control (n=114) and PCOS (n=102) subjects.

| Sl No | Cytokines | Control (mean± SEM) | PCOS (mean± SEM) | P value |
| --- | --- | --- | --- | --- |
| 1 | IL18 | 26.7 ± 2.2 | 50.1 ± 8.6 | <0.01 |
| 2 | IL-1Ra | 394.5 ± 38.3 | 701.9 ± 80.2 | <0.001 |
| 3 | IL12p70 | 70.1 ± 6.6 | 18.9 ± 8.7 | <0.05 |
| 4 | IL13 | 2.1 ± 0.3 | 8.7 ± 3.1 | <0.05 |
| 5 | IL16 | 155.9 ± 25.4 | 257.8 ± 27.2 | <0.01 |
| 6 | MCP-1(CCL2) | 23.9 ± 1.9 | 42.9 ± 3.9 | <0.0001 |
| 7 | MCP-3(CCL7) | 0.82 ± 0.1 | 4.3 ± 1.8 | <0.05 |
| 8 | CTACK(CCL27) | 362.1 ± 23.4 | 484.4 ± 33.6 | <0.01 |
| 9 | Eotaxin(CCL11) | 48.5 ± 3.7 | 65.6 ± 6.2 | <0.05 |
| 10 | RANTES(CCL5) | 17381.3 ± 1299.2 | 50052.6 ± 12952.4 | <0.01 |
| 11 | IP-10(CXCL-10) | 276.8 ± 21.8 | 401.4 ± 44.7 | <0.05 |
| 12 | M-CSF(CSF-1) | 14.7 ± 1.2 | 26.9 ± 4.3 | <0.01 |
| 13 | HGF | 320.7 ± 25.9 | 450.4 ± 57.8 | <0.05 |
| 14 | β-NGF | 5.4 ± 0.9 | 18.6 ± 6.9 | <0.05 |

**Footnote for Table 2:** All values were presented as mean± SEM. The concentration of cytokines is expressed in pg/mL.

Additionally, the relative differential expression of significantly altered 14 plasma cytokines was analyzed in four PMOS phenotypes compared with the control subjects. Our comparative analysis revealed there is a significant difference in the cytokine levels of IL-1Ra (p-value<0.01), IL16 (p-value<0.05), MCP-1 (p-value<0.001), CTACK (p-value<0.0001), Eotaxin (p-value<0.05), RANTES (p-value<0.01), and IP-10 (p-value<0.01) between the control and PMOS phenotype A groups. Also, a significant difference was observed in IL18 (p-value<0.001), IL-1Ra (p-value<0.05), IL12p70 (p-value<0.0001), IL13 (p-value <0.0001), MCP-1 (p-value<0.001), MCP-3 (p-value<0.0001), M-CSF (p-value<0.0001), HGF (p-value<0.0001) and β-NGF (p-value<0.0001) between control and phenotype B groups. Whereas, even with the second highest population, phenotype C and phenotype D didn’t show any significant change compared to control groups (Figure 3). The concentration of 14 altered cytokines in each PMOS phenotype is given in the Supplementary Table S2.

**Figure 3:**
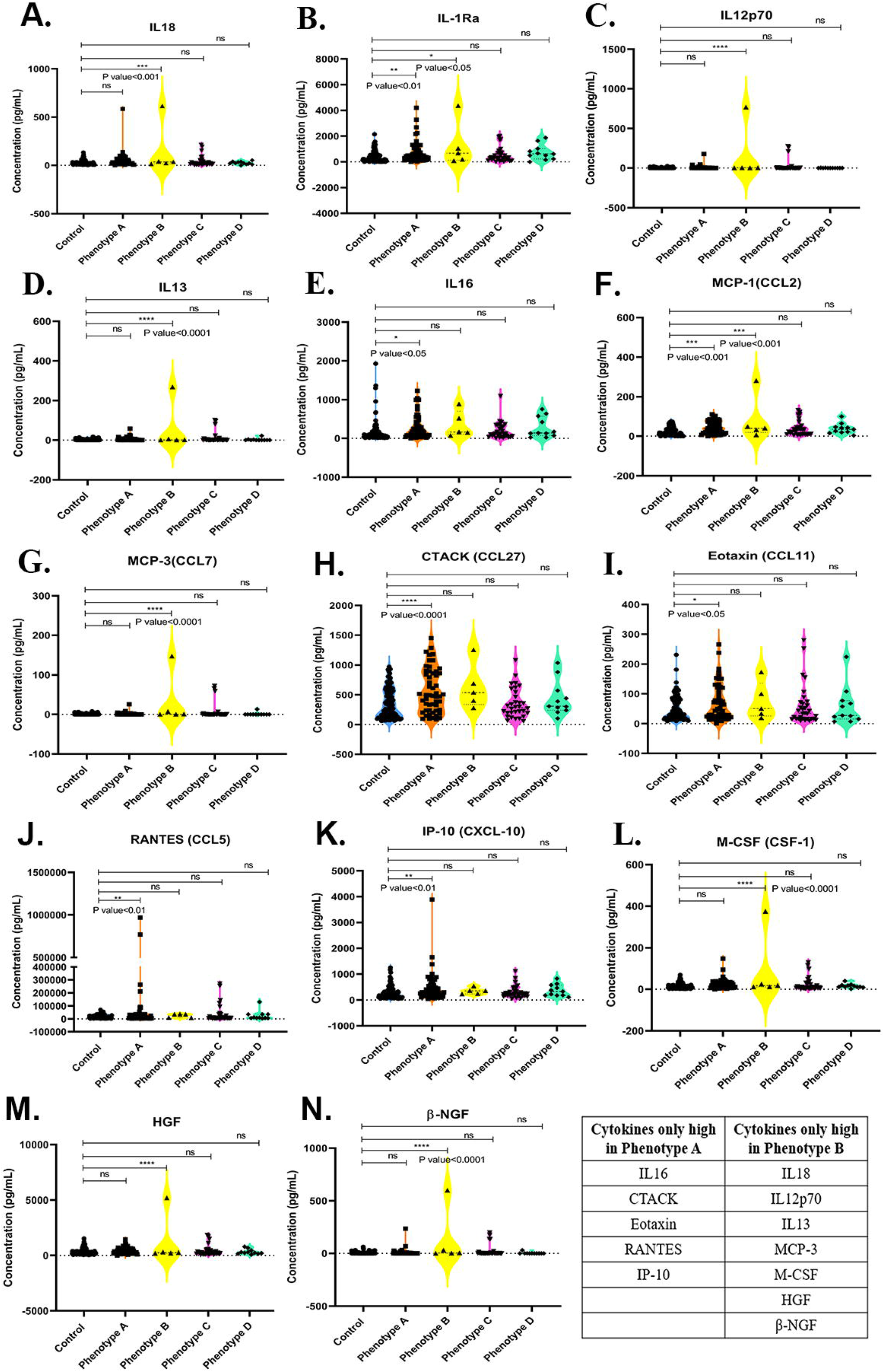
Comparative analysis of various significantly altered cytokines in control and four PMOS phenotypes. Comparison of mean cytokine concentration (pg/mL) change in control and four different PMOS phenotypes: phenotype A (HA+OD+PCOM), phenotype B (HA+OD), phenotype C (HA+PCOM) and phenotype D (OD+PCOM). Graphs are depicted as the violin with a dot plot representation of the significantly altered cytokines. Blue colour represents control, orange colour represents phenotype A, yellow colour represents phenotype B, pink colour represents phenotype C and green colour represents phenotype D. Each dot is represented as a single sample, and (p-value < 0.05) is considered statistically significant.

### 2.3. ROC analysis showed significant diagnostic potential of altered cytokines in PMOS condition

We have performed ROC analysis for 12 cytokines specifically altered in phenotype A and phenotype B. The ROC analysis for the other two cytokines (IL-1RA and MCP1) were not performed as they were altered in both the phenotypes. A total of 5 cytokines specifically altered in phenotype A and 7 cytokines specifically altered in phenotype B were used for the analysis. The ROC analysis revealed a panel of three cytokines (IL16, CTACK, and IP-10) with a AUC value >0.6 shows a higher diagnostic potential for PMOS phenotype A diagnosis. Whereas, another panel of three cytokines (IL18, M-CSF, and HGF) with AUC value>0.6 revealed their significant diagnostic potential for PMOS phenotype B diagnosis (Figure 4).

**Figure 4:**
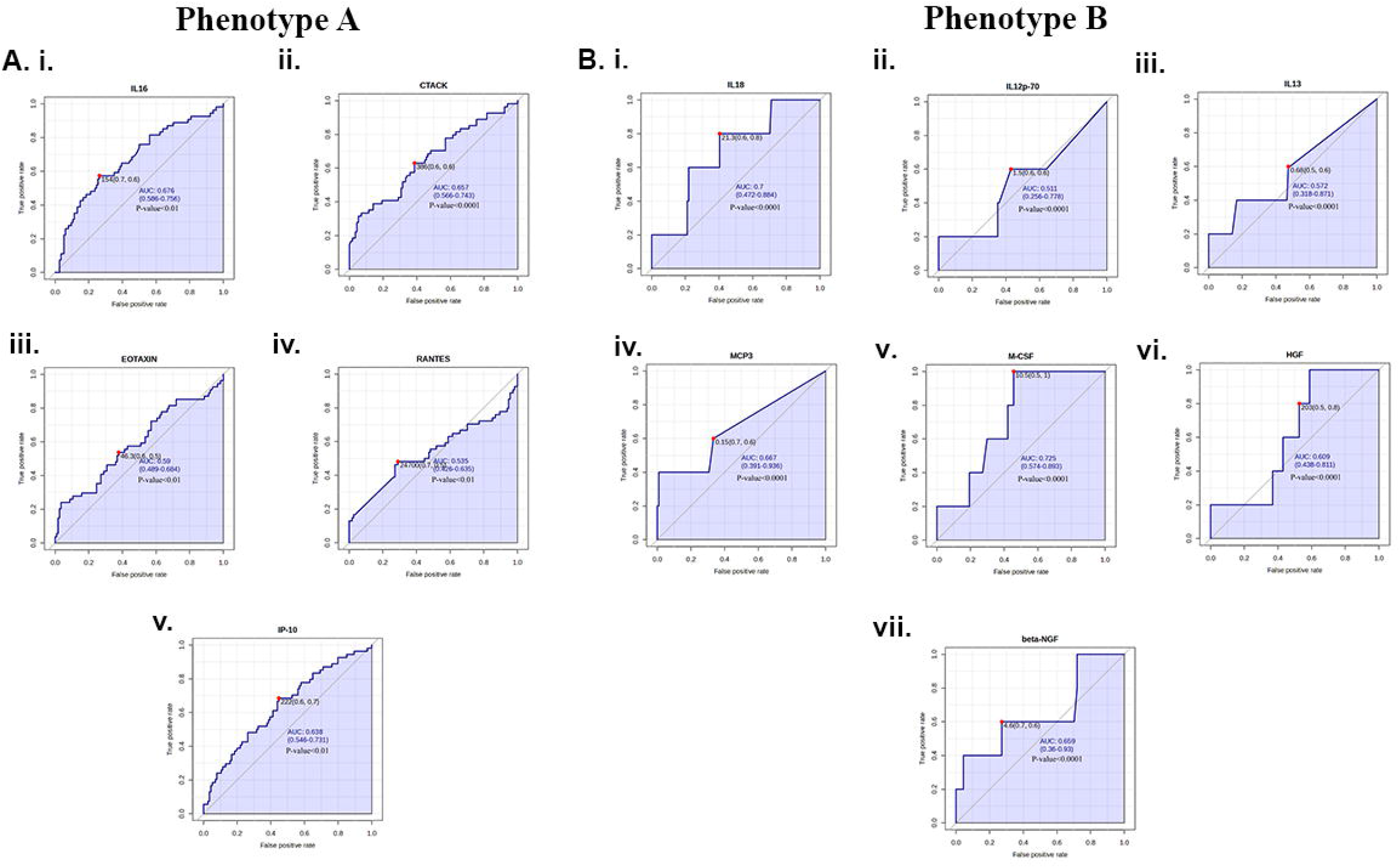
ROC analysis of altered cytokines in specific to PMOS Phenotype A and B. ROC analysis of 5 cytokines specifically altered in phenotype A and 7 cytokines altered in phenotype B. Graphs A(i-v) shows ROC curves of IL16, CTACK, EOTAXIN, RANTES, and IP-10. Graphs B(i-vii) shows ROC curves of IL18, IL12p-70, IL13, MCP3, M-CSF, HGF, and beta-NGF. Each graph is represented with its AUC value, class interval, and t-test value. X-axis shows the False positive rate, while Y-axis shows the True positive rate.

### 2.4. Odds ratio of altered cytokines revealed their potential for PMOS developement

To further assess the relationship between altered cytokine levels and PMOS susceptibility, we evaluated the odds ratio (OR) of significantly altered cytokines (Supplementary Table S3). Our OR analysis identified eleven cytokines and growth factors exhibited OR values greater >1 with IL-16 highest OR; MCP-1 (CCL2), M-CSF, IL-1Ra (OR range of 2 to 3) and IL-18, IL-13, CTACK (CCL27), Eotaxin (CCL11), RANTES (CCL5), IP-10 (CXCL10), and HGF (OR range of 1 to 2).

Thus, signifying a correlation of these markers with the risk of getting PMOS and hence exhibiting a diagnostic value as shown in Figure 5.

**Figure 5:**
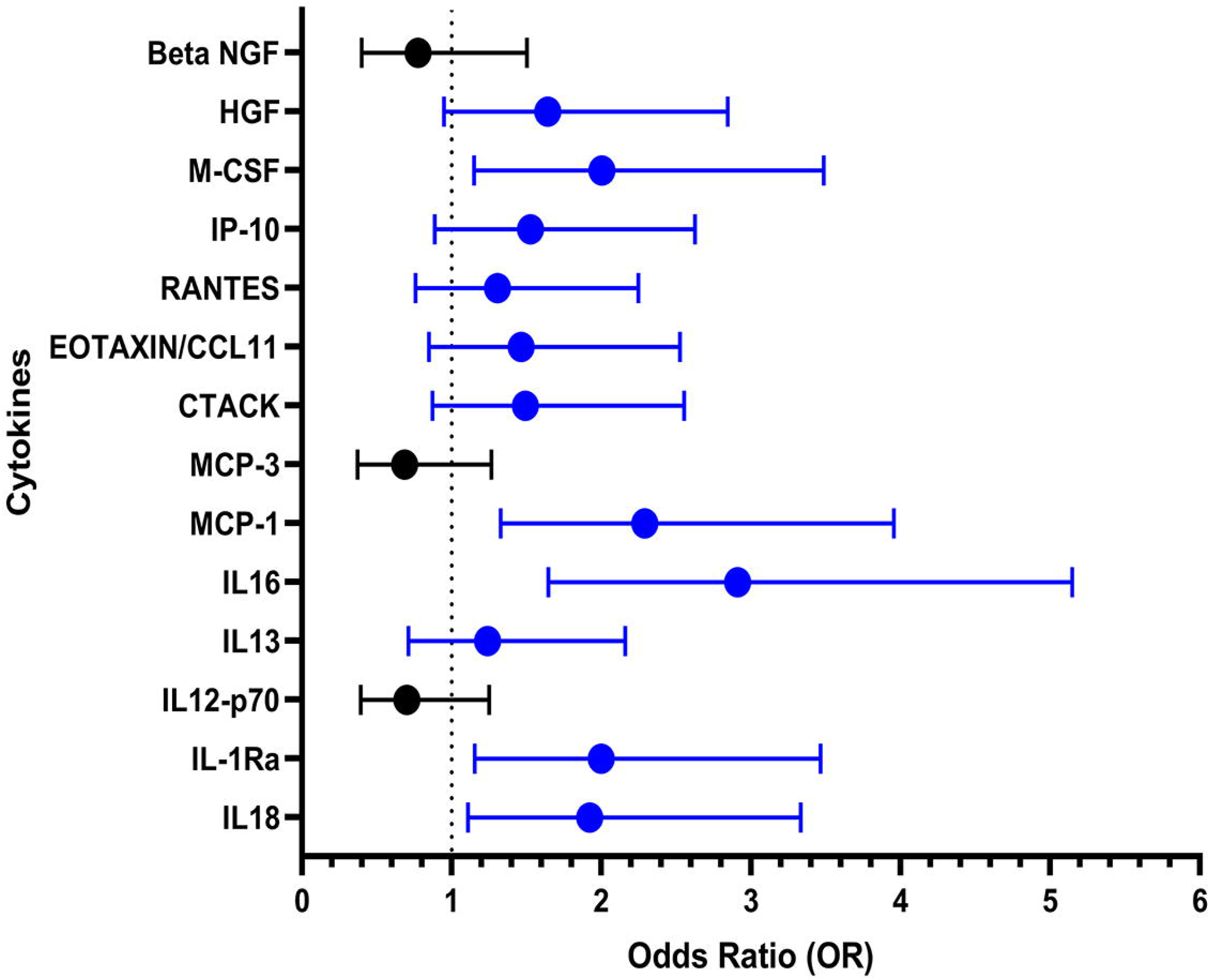
Odds ratio calculation representing a correlation between cytokines and the risk of PMOS. Odds ratio analysis of altered cytokines. All values were presented in the Forest plot as lower 95% CI, odds ratio, and upper 95% CI. Cytokines with OR >1 (coloured Blue) signify a stronger correlation with the risk of getting PMOS.

### 2.5. Correlation analysis of altered cytokines with each other in control and PMOS condition

To identify the correlation of each cytokine to the other, we have performed Pearson’s correlation coefficient calculations for all possible pairwise combinations between each 11 significantly altered cytokines (OR>1). Entirely, the calculation was shown for a total of 121 pairwise correlation combinations in each correlation heatmap matrix, as shown in Figure 6. In control subjects, we observed 108 significant positive correlations, among which 8 pairs including IL16-Eotaxin, IL18-MCSF, IL18-HGF, MCP1-CTACK, MCP1-Eotaxin, CTACK-Eotaxin, CTACK-HGF and IP10-HGF. While, in the PMOS condition, we observed 84 significant positive correlations. Out of which, 10 combinations of cytokines IL16-IL1Ra, IL18-IL13, IL18-MCSF, IL18-HGF, IL13-MCP-1, IL13-MCSF, IL13-HGF, MCP1-MCSF, MCP1-HGF, MCSF-HGF and vice versa; showed stronger positive correlation with R-value ≥ 0.7 (Supplementary Table S4). In the correlation matrix of PMOS, a total of 2 pairwise combinations (out of 121 combinations), such as (IL13-RANTES (R= −0.06) and vice versa) showed a stronger negative correlation (R-value ≤ −0.7), but are not statistically significant. Hence, not considered in our study. It is interesting to note that IL18-IL13, IL13-MCP-1, IL13-MCSF, IL13-HGF, IL16-IL1Ra, MCP1-MCSF, MCP1-HGF and MCSF-HGF pairs show specific correlation in the diseased state while IL18-MCSF and IL18-HGF pair exhibit correlation in both states suggesting the importance of these 8 cytokine pairs in PMOS pathogenesis. In contrast, the strong IP10-HGF correlation in the normal state is switched to the HGF-MCSF correlation in the pathological state of PMOS. On the other hand, we also observed that most of the cytokines were moderately correlated (R-value-0.5 to 0.7) in control women. Whereas, in the PMOS condition few cytokines showed significantly stronger correlation by forming clusters and few moderately correlated cytokines lost their correlation. This signifies an optimum balance in cytokine signaling in normal women, while an aberrant cytokine signaling alters immune etiology in PMOS conditions.

**Figure 6:**
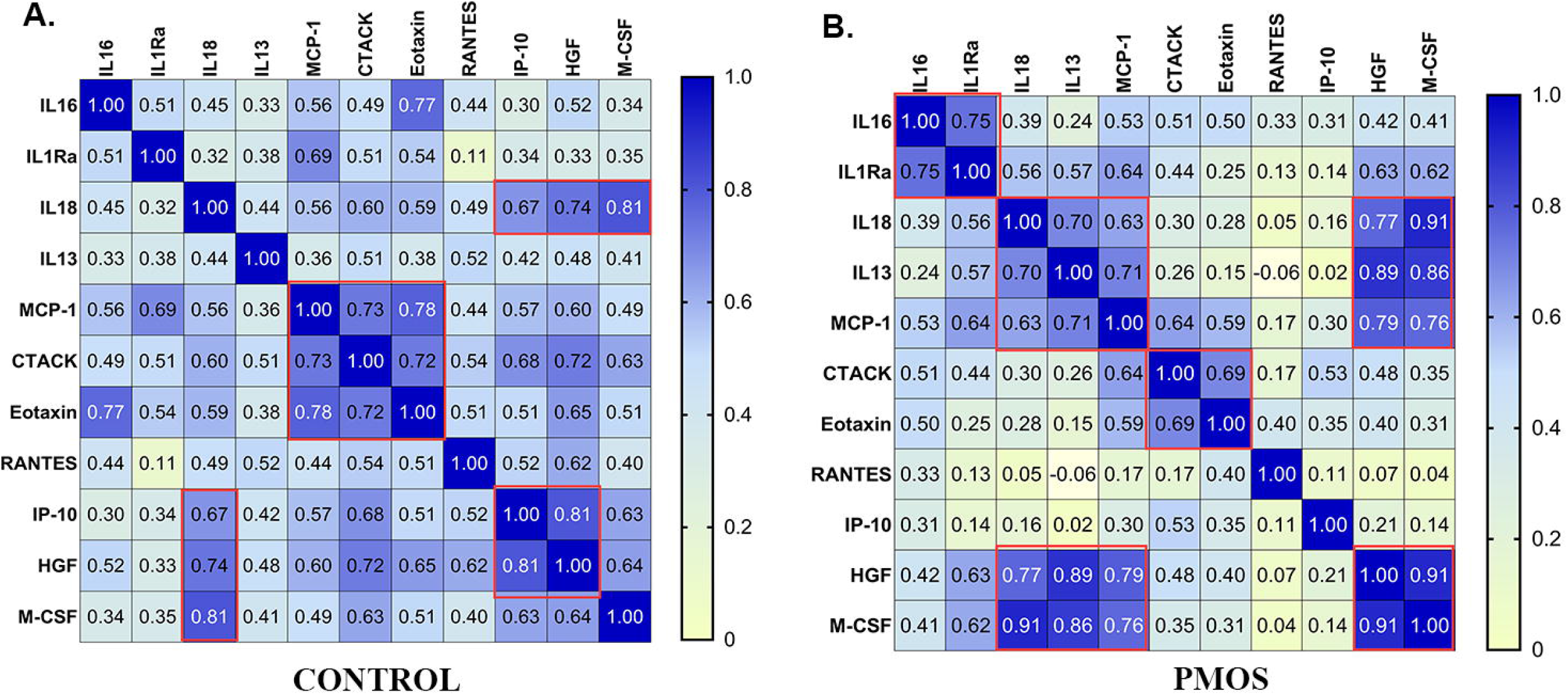
Pearson’s correlation heat map showing the correlation between various altered cytokines in control and PMOS condition. Correlation matrix of altered cytokines (OR>1) in control and PMOS subjects: (A) Control and (B) case or PMOS. Heatmap with R values in relative boxes summarizes the results of Pearson’s correlation analysis between the cytokines. Positive correlations were represented as shadows of blue and negative correlations were represented as shadows of yellow.

### 2.6. GOnet network analysis

To understand the networks and biological processes in which these 11 high diagnostic valued cytokines (with OR>1) were involved, we have performed GOnet network biological process analysis. Enrichment analysis ≤ 0.05 was considered for our network selection and unconnected or poorly connected networks were removed from our analysis. The specific enriched networks (Figure 7B, Supplementary Data File S1) involved strongly interacted cytokines: CXCL10 which is associated with inflammatory and immune response, response to cytokine/chemokine stimuli, response to external stimuli (chemical, biotic, lipopolysaccharide, molecules of bacterial origin, lipid, organic substances), regulation of chemotaxis (including leukocyte, lymphocyte, T-cell, eosinophil, neutrophil, granulocyte, monocyte, natural killer cells), positive regulation of metabolic process; IL13 and CCL5 for regulation of protein phosphorylation, positive regulation to JAK-STAT pathway and STAT signaling, inflammatory process, leukocytes, and lymphocytes proliferation and activation. The moderately interacted cytokines: IL-1RN, CCL2, IL18, and IL16 were involved in processes such as regulation of protein phosphorylation, T-cell and neutrophil chemotaxis, MAPK cascade, Tyrosine phosphorylation of STAT and neutrophil migration as shown in Figure 7A and 7B. Further, phenotype A and phenotype B-specific Gonet network enrichment analysis was performed separately (Figures 7C and 7D). Cytokines specifically significantly upregulated in phenotype A such as CCL5, IL16, CXCL10, CCL11, and CCL27 were mainly involved in mononuclear cells, T cells, and neutrophils migration and chemotaxis, regulation of cell motility, cellular and sub-cellular component movement, and inflammatory response. Whereas cytokines specifically significantly upregulated in phenotype B such as IL13, CSF1, IL18, and HGF were mainly involved in mononuclear cell and leukocyte proliferation, macrophage-derived foam cell differentiation, regulation to protein tyrosine phosphorylation, regulation of JAK-STAT pathway and phosphorylation of STAT and regulation of phosphatidylinositol 3-kinase signaling. This signifies that PMOS phenotype A is strongly associated with aberrant chemotaxis and immune cell migration that may lead to altered ovarian immune micro-environment and resulting PCO morphology. While phenotype B is strongly associated with altered immune cell signaling pathways.

**Figure 7:**
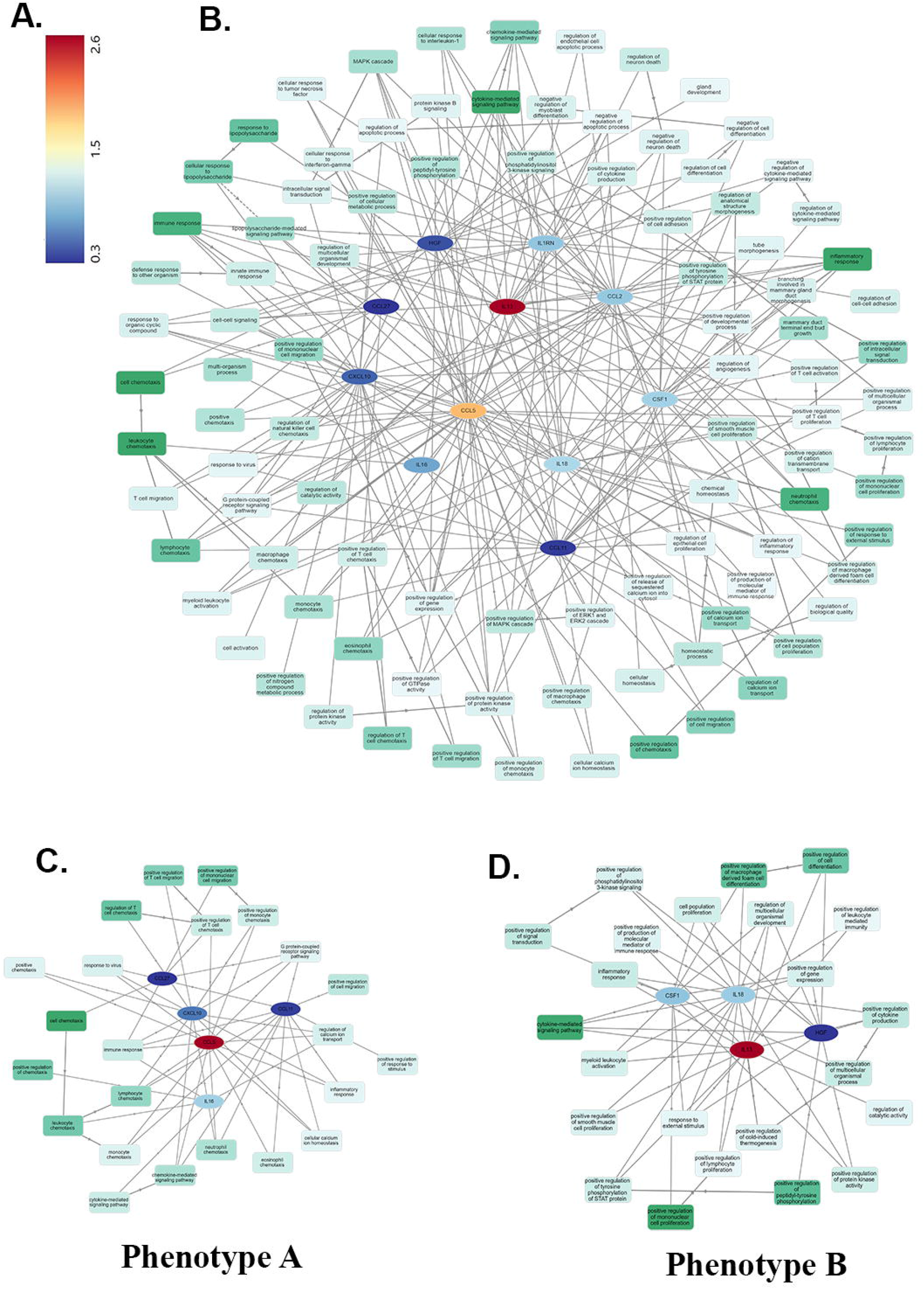
GOnet network analysis of altered cytokines. GOnet network analysis of altered cytokines. (A) Colour matrix of blue to red (left corner) showed along with FC, (B) GOnet network of 11 significantly altered cytokines (with OR>1), (C) GOnet network of cytokines only high in phenotype A, (D) GOnet network of cytokines only high in phenotype B. The strongly involved cytokines with a higher fold change value were represented as red colour shadows and weakly involved cytokines with a lower fold change value were in blue colour shadows (oval shape). Similarly, the strongly involved networks are shown in shadows of dark green (square shape).

## 3. Discussion

PMOS is increasingly recognized as a chronic low-grade inflammatory disorder characterized by immune dysregulation, hyperandrogenism, insulin resistance, and obesity. This inflammatory milieu promotes steroidogenic enzyme expression and androgen production.(2, 14). In this study, profiling of 48 circulating cytokines identified 14 significantly altered cytokines, including IL-18, IL-16, IL-12p70, IL-13, IL-1Ra, MCP-1, MCP-3, CTACK, Eotaxin, RANTES, IP-10, M-CSF, HGF, and β-NGF, supporting systemic inflammation and immune dysregulation in PMOS.

Among the altered cytokines, IL-18 and IL-16 emerged as key inflammatory mediators. Elevated IL-18 confirms previous reports linking it to hyperandrogenism, independent of insulin resistance or obesity(15, 16) and may also explain the increased IgE levels observed in PMOS through its role in IgE accumulation(17).

IL-16 showed the highest odds ratio among all cytokines, indicating a strong association with disease risk. Given their ability to activate the p38 MAPK pathway (18), elevated IL-18 and IL-16 may be responsible for the overactivated p38MAPK pathway in PMOS (19) and contribute to chronic inflammation in PMOS, potentially through a feedback loop between IL-16 and MAPK signaling(20).

We observed significantly elevated plasma IL-13 levels in PMOS, consistent with previous reports of increased IL-13 in follicular fluid(21). In contrast, IL-15 and IFN-γ levels were unchanged. As IL-18 induces IFN-γ only in the presence of IL-15, its elevation in the absence of IL-15 may instead promote Th2 differentiation and IL-13 production(22). These findings suggest a shift toward Th2-mediated immune responses that may contribute to T-cell dysregulation in PMOS(23).

IL-12p70, a key Th1-polarizing cytokine that drives naïve CD4⁺ T-cell differentiation into Th1 cells, was significantly elevated in the plasma of PMOS patients. This contrasts with previous reports of reduced IL-12 in follicular fluid(21), likely reflecting differences between systemic and ovarian immune environments. Further studies comparing cytokine profiles across biological compartments are needed to clarify these differences.

Chemokines regulate immune cell migration during inflammation. We observed significantly elevated plasma MCP-1 (CCL2) levels in PMOS, consistent with its role in monocyte recruitment and chronic inflammation(24). As MCP-1 is linked to obesity (25), atherosclerosis and cardiometabolic disease(26), its elevation may contribute to the chronic inflammatory state and increased cardiovascular risk in PMOS. Increased MCP-1 expression may also be partly explained by promoter polymorphisms in the **MCP1** gene(27).

PMOS is frequently associated with insulin resistance and inflammatory skin disorders. We observed significantly elevated plasma levels of CTACK (CCL27), Eotaxin (CCL11), and RANTES (CCL5), chemokines implicated in psoriasis(28, 29) atopic dermatitis(30–32), and other inflammatory skin diseases(33). Their concurrent elevation suggests enhanced recruitment of T cells, eosinophils, and macrophages to the skin, potentially contributing to the dermatological manifestations of PMOS. Elevated IL-13 may further promote Eotaxin expression through p38 MAPK-dependent signaling(38), supporting a possible p38 MAPK–IL-13–CCL11 inflammatory axis in PMOS. RANTES, also linked to metabolic syndrome and type 2 diabetes (34, 35), may contribute to both systemic inflammation and cardiometabolic risk. Additionally, increased IL-16 and RANTES levels indicate enhanced recruitment of dendritic cells(36, 37)and CD4⁺ T cells (38), supporting persistent immune activation and dysregulated adaptive immune responses in PMOS.

A significant increase in circulating IP-10 (CXCL10) was observed in PMOS, consistent with previous reports (39). IP-10 recruits NK cells, monocytes, macrophages, dendritic cells, and activated T lymphocytes and has been linked to obesity(40), type 2 diabetes(41) and cardiovascular disease(42), suggesting a role in inflammation-driven metabolic dysfunction. As IL-18 can induce both IP-10 (43) and and RANTES expression(CCL5)(44), their concurrent elevation indicates activation of a coordinated pro-inflammatory chemokine network in PMOS. Together with increased IL-16, MCP-1, and CTACK, these findings further support persistent low-grade inflammation and immune activation underlying the metabolic and reproductive abnormalities of PMOS.

We observed significantly elevated plasma levels of IL-1 receptor antagonist (IL-1Ra) in women with PMOS. IL-1Ra is an anti-inflammatory cytokine that counteracts IL-1 signaling and is secreted by adipose tissue in response to metabolic stress(45, 46). Its levels correlate with BMI and are increased in overweight and obese women with PMOS(47). Although IL-1Ra likely represents a compensatory response to chronic inflammation, persistently elevated levels have also been associated with impaired insulin secretion, insulin resistance(47), and metabolic dysfunction, reflecting the severity of metabolic disturbances in PMOS.

We observed significantly elevated plasma M-CSF (CSF-1) levels in PMOS. M-CSF regulates monocyte–macrophage proliferation, survival, and activation, thereby promoting inflammatory responses. Consistent with previous reports of increased CSF1 expression in the PMOS endometrium(48), elevated M-CSF may also contribute to increased IL-18 production(49), amplifying inflammatory signaling. In addition, HGF and β-NGF levels were significantly increased, suggesting that altered growth factor signaling, alongside immune dysregulation, may contribute to tissue remodeling, angiogenesis, ovarian function, and the metabolic and reproductive abnormalities of PMOS.

Comparative analysis of the four PMOS phenotypes revealed significant cytokine and chemokine alterations primarily in phenotypes A and B, whereas phenotypes C and D showed no significant changes, highlighting the immunometabolic heterogeneity of PMOS. Phenotype A exhibited elevated IL-1Ra, MCP-1, and RANTES levels together with higher BMI, consistent with its association with obesity(45, 47), insulin resistance, and increased cardiometabolic risk(50). Phenotype B showed increased IL-18, MCP-1, and M-CSF, indicating higher prevalence of obesity and hypertriglyceridemia(50), insulin resistance and obesity(15) and enhanced monocyte/macrophage activation and chronic inflammation. These findings suggest that inflammatory burden is greatest in the metabolically compromised phenotypes A and B, while phenotypes C and D display comparatively milder immune dysregulation.

We identified inflammatory mediators with potential diagnostic value for phenotype-specific PMOS classification. ROC analysis showed that IL-16, CTACK (CCL27), and IP-10 (CXCL10) best discriminated phenotype A, whereas IL-18, M-CSF, and HGF were most informative for phenotype B, suggesting distinct inflammatory signatures across PMOS phenotypes. Odds ratio analysis identified IL-16, followed by MCP-1, M-CSF, and IL-1Ra, as the strongest disease-associated biomarkers. Correlation analysis further revealed a highly interconnected inflammatory network involving interleukins, chemokines, M-CSF, and HGF, supporting coordinated immune activation underlying the chronic low-grade inflammation characteristic of PMOS.

Gene Ontology network (GOnet) analysis demonstrated that the altered cytokines were enriched in immune pathways regulating leukocyte, neutrophil, and T-cell chemotaxis, inflammatory responses, cell migration, and protein phosphorylation, indicating coordinated immune-cell recruitment and activation in PMOS. Phenotype-specific analysis revealed enhanced chemotaxis-related pathways in phenotype A and intracellular signaling pathways, including PI3K signaling and tyrosine phosphorylation, in phenotype B, highlighting distinct immunological mechanisms. Importantly, cytokine alterations were independent of BMI, suggesting they are intrinsic to PMOS rather than secondary to obesity. Although follicular fluid cytokine profiling was not performed, our findings identify coordinated immune networks and phenotype-specific inflammatory signatures that support immune dysregulation as a central feature of PMOS and highlight potential biomarkers and therapeutic targets.

## Significance Statement

This study provides a comprehensive profile of 48 circulating cytokines in women with PMOS, identifying 14 elevated inflammatory mediators and phenotype-associated cytokine signatures (Graphical Abstract). The prominence of IL-16 and the enrichment of chemotaxis and immune cell migration pathways point to potential mechanisms underlying systemic immune dysregulation in PMOS. These findings advance our understanding of the immunological heterogeneity of PMOS and provide a foundation for validating circulating cytokines as candidate biomarkers for disease characterization and phenotype stratification.

## Acknowledgements

We acknowledge the support of BIORAD in conducting the Bio-Plex Pro^Tm^ Human Cytokine Screening Assay. The support extended by KJK Hospital and SAMAD IVF Hospital staff is also acknowledged for patient recruitment and data management.

## Data Availability Statement

The data for the study essential to this article are available in the article itself and its online Supplementary material. However, the data underlying Table 1 of this article cannot be shared publicly due to the privacy of individuals who participated in the study. The data will be shared upon reasonable request to the corresponding author.

## Authors’ Contribution

ML contributed to project conception, funding acquisition, study design, project administration supervision, and manuscript review and editing. LPP executed all the experiments, while BSJ executed experiments pertaining to Figure 2 and Figure S1, and both contributed to the original manuscript draft. LPP analyzed and interpreted all the data for Figures 1-7 and all supplementary figures. The diagnosis and recruitment of PMOS patients were done by KJK and SMP. All authors were involved in the discussion of the results and approved the final article.

## Glossary/Abbreviations

PMOS (Polycystic ovary syndrome), IR (Insulin resistance), EDTA (Ethylenediaminetetraacetic acid), NaCl (Sodium Chloride), RBC (Red blood corpuscles), ELISA (enzyme-linked immunosorbent assay), LH (Leutenizing hormone), FSH (Follicle stimulating hormone), Basic FGF (basic fibroblast growth factor), CTACK (cutaneous T-cell attracting chemokine), G-CSF (granulocyte colony-stimulating factor), GM-CSF (granulocyte-macrophage colony-stimulating factor), GRO-α (growth-regulated oncogene α), HGF (hepatocyte growth factor), IFNα2 (interferon α2), IFNγ (interferon γ), IL1α (Interleukin 1α), IL1β (Interleukin 1β), IL-1Ra (Interleukin - 1receptor antagonist), IL2 (Interleukin 2), IL-2Rα (Interleukin-2 receptor α), IL3 (Interleukin 3), IL4 (Interleukin 4), IL5 (Interleukin 5), IL6 (Interleukin 6), IL7 (Interleukin 7), IL8 (Interleukin 8), IL9 (Interleukin 9), IL10 (Interleukin 10), IL12p40 (Interleukin 12 p40, IL12p70 (Interleukin 12p70), IL13 (Interleukin 13), IL15 (Interleukin 15), IL16 (Interleukin 16), IL17A (Interleukin 17A), IL18(Interleukin 18), IP-10 (Interferon gamma-induced protein 10), LIF (leukemia inhibitory factor), MCP-1(monocyte chemotactic protein 1), MCP-3 (monocyte chemotactic protein 3), M-CSF (macrophage colony-stimulating factor), MIF (macrophage migration inhibitory factor), MIG (Monokine induced by IFNγ), (MIP-1α (macrophage inflammatory protein 1α), MIP-1β (macrophage inflammatory protein 1β), β-NGF (β-nerve growth factor), PDGF-BB (platelet-derived growth factor BB), RANTES (Regulated on activation normal T cell expressed and secreted), SCF (Stem cell factor), SCGF-β (Stem cell growth factor β), SDF-1α (Stromal cell– derived factor 1α), TNFα (Tumor necrosis factor α), TNFβ (Tumor necrosis factor β), TRAIL (TNF-related apoptosis inducing ligand), VEGF (vascular endothelial growth factor), RT (Room temperature), OR (Odds ratio), CI (Class interval), ROC (Receiver Operator Characteristics), BMI (body mass index), SD (standard deviation), CCL2 (chemokine (C-C motif) ligand 2), CCL7 (chemokine (C-C motif) ligand 7), CCL11 (chemokine (C-C motif) ligand 11), CCL27 (chemokine (C-C motif) ligand 27), CCL5 (chemokine (C-C motif) ligand 5), CXCL-10 (C-X-C motif chemokine ligand 10), CSF-1(Colony stimulating factor-1), CSF-2(Colony stimulating factor-2), AUC (Area under curve), LCF (lymphocyte chemoattractant factor), DC (dendritic cell).

## References

1. Sirmans SM, Pate KA. Epidemiology, diagnosis, and management of polycystic ovary syndrome. Clinical epidemiology. 2013:1–13.

2. Zangeneh FZ, Naghizadeh MM, Masoumi M. Polycystic ovary syndrome and circulating inflammatory markers. International Journal of Reproductive BioMedicine. 2017;15(6):375.

3. Dunaif A. Insulin resistance and the polycystic ovary syndrome: mechanism and implications for pathogenesis. Endocrine reviews. 1997;18(6):774–800.

4. Kelly CC, Lyall H, Petrie JR, Gould GW, Connell JM, Sattar N. Low grade chronic inflammation in women with polycystic ovarian syndrome. The Journal of Clinical Endocrinology & Metabolism. 2001;86(6):2453–5.

5. Kalyan S, Goshtesabi A, Sarray S, Joannou A, Almawi WY. Assessing C reactive protein/albumin ratio as a new biomarker for polycystic ovary syndrome: a case–control study of women from Bahraini medical clinics. BMJ open. 2018;8(10):e021860.

6. Orio Jr F, Palomba S, Cascella T, Di Biase S, Manguso F, Tauchmanovà L, et al. The increase of leukocytes as a new putative marker of low-grade chronic inflammation and early cardiovascular risk in polycystic ovary syndrome. The Journal of Clinical Endocrinology & Metabolism. 2005;90(1):2–5.

7. Repaci A, Gambineri A, Pasquali R. The role of low-grade inflammation in the polycystic ovary syndrome. Molecular and cellular endocrinology. 2011;335(1):30–41.

8. Bornstein S, Rutkowski H, Vrezas I. Cytokines and steroidogenesis. Molecular and cellular endocrinology. 2004;215(1-2):135–41.

9. Büscher U, Chen F, Kentenich H, Schmiady H. Cytokines in the follicular fluid of stimulated and non-stimulated human ovaries; is ovulation a suppressed inflammatory reaction? Human reproduction. 1999;14(1):162–6.

10. Evans J, Salamonsen LA. Inflammation, leukocytes and menstruation. Reviews in Endocrine and Metabolic Disorders. 2012;13:277–88.

11. Verthelyi D, Klinman D. Sex hormone levels correlate with the activity of cytokine-secreting cells in vivo. Immunology. 2000;100(3):384–90.

12. Möhlig M, Spranger J, Osterhoff M, Ristow M, Pfeiffer A, Schill T, et al. The polycystic ovary syndrome per se is not associated with increased chronic inflammation. European Journal of Endocrinology. 2004;150(4):525–32.

13. Azziz R, Carmina E, Dewailly D, Diamanti-Kandarakis E, Escobar-Morreale HF, Futterweit W, et al. The Androgen Excess and PCOS Society criteria for the polycystic ovary syndrome: the complete task force report. Fertility and sterility. 2009;91(2):456–88.

14. Shorakae S, Teede H, de Courten B, Lambert G, Boyle J, Moran LJ, editors. The emerging role of chronic low-grade inflammation in the pathophysiology of polycystic ovary syndrome. Seminars in reproductive medicine; 2015: Thieme Medical Publishers.

15. Escobar-Morreale HcF, Botella-Carretero JI, Villuendas G, Sancho J, San Millán JL. Serum interleukin-18 concentrations are increased in the polycystic ovary syndrome: relationship to insulin resistance and to obesity. The journal of clinical endocrinology & metabolism. 2004;89(2):806–11.

16. Yang Y, Qiao J, Li R, Li M-Z. Is interleukin-18 associated with polycystic ovary syndrome? Reproductive Biology and Endocrinology. 2011;9:1–5.

17. Yoshimoto T, Mizutani H, Tsutsui H, Noben-Trauth N, Yamanaka K-i, Tanaka M, et al. IL-18 induction of IgE: dependence on CD4+ T cells, IL-4 and STAT6. Nature immunology. 2000;1(2):132–7.

18. Krautwald S. IL-16 activates the SAPK signaling pathway in CD4+ macrophages. The Journal of Immunology. 1998;160(12):5874–9.

19. Krishna MB, Johnson BS, Vasudevan M, Pillai SM, Laloraya M. miRNA-mRNA Network in PBMCs of PCOS Women Identifies Overactivated Stress-Activated Kinases. Cellular Physiology & Biochemistry (Cell Physiol Biochem Press GmbH & Co KG). 2023;57(2).

20. Laurence A, Astoul E, Hanrahan S, Totty N, Cantrell D. Identification of pro-interleukin 16 as a novel target of MAP kinases in activated T lymphocytes. European journal of immunology. 2004;34(2):587–97.

21. Gallinelli A, Ciaccio I, Giannella L, Salvatori M, Marsella T, Volpe A. Correlations between concentrations of interleukin-12 and interleukin-13 and lymphocyte subsets in the follicular fluid of women with and without polycystic ovary syndrome. Fertility and sterility. 2003;79(6):1365–72.

22. Hoshino T, Wiltrout RH, Young HA. IL-18 is a potent coinducer of IL-13 in NK and T cells: a new potential role for IL-18 in modulating the immune response. The Journal of Immunology. 1999;162(9):5070–7.

23. Krishna MB, Joseph A, Subramaniam AG, Gupta A, Pillai SM, Laloraya M. Reduced Tregs in peripheral blood of PCOS patients–a consequence of aberrant Il2 signaling. The Journal of Clinical Endocrinology & Metabolism. 2015;100(1):282–92.

24. Deshmane SL, Kremlev S, Amini S, Sawaya BE. Monocyte chemoattractant protein-1 (MCP-1): an overview. Journal of interferon & cytokine research. 2009;29(6):313–26.

25. Takahashi K, Mizuarai S, Araki H, Mashiko S, Ishihara A, Kanatani A, et al. Adiposity elevates plasma MCP-1 levels leading to the increased CD11b-positive monocytes in mice. Journal of Biological Chemistry. 2003;278(47):46654–60.

26. Deo R, Khera A, McGuire DK, Murphy SA, de P. Meo Neto J, Morrow DA, et al. Association among plasma levels of monocyte chemoattractant protein-1, traditional cardiovascular risk factors, and subclinical atherosclerosis. Journal of the American College of Cardiology. 2004;44(9):1812–8.

27. Vasyukova E, Zaikova E, Kalinina O, Gorelova I, Pyanova I, Bogatyreva E, et al. Inflammatory and anti-inflammatory parameters in PCOS patients depending on body mass index: A case-control study. Biomedicines. 2023;11(10):2791.

28. Campanati A, Goteri G, Simonetti O, Ganzetti G, Giuliodori K, Stramazzotti D, et al. CTACK/CCL27 expression in psoriatic skin and its modification after administration of etanercept. British Journal of Dermatology. 2007;157(6):1155–60.

29. Kakinuma T, Saeki H, Tsunemi Y, Fujita H, Asano N, Mitsui H, et al. Increased serum cutaneous T cell-attracting chemokine (CCL27) levels in patients with atopic dermatitis and psoriasis vulgaris. Journal of allergy and clinical immunology. 2003;111(3):592–7.

30. Holm JG, Hurault G, Agner T, Clausen ML, Kezic S, Tanaka RJ, et al. Immunoinflammatory biomarkers in serum are associated with disease severity in atopic dermatitis. Dermatology. 2021;237(4):513–20.

31. Yawalkar N, Uguccioni M, Schärer J, Braunwalder J, Karlen S, Dewald B, et al. Enhanced expression of eotaxin and CCR3 in atopic dermatitis. Journal of investigative dermatology. 1999;113(1):43–8.

32. Niwa Y. Elevated RANTES levels in plasma or skin and decreased plasma IL-10 levels in subsets of patients with severe atopic dermatitis. Archives of dermatology. 2000;136(1):125–6.

33. Amerio P, Frezzolini A, Feliciani C, Verdolini R, Teofoli P, Pit O, et al. Eotaxins and CCR3 receptor in inflammatory and allergic skin diseases: therapeutical implications. Current Drug Targets-Inflammation & Allergy. 2003;2(1):81–94.

34. Herder C, Haastert B, Muller-Scholze S, Koenig W, Thorand B, Holle R, et al. Association of systemic chemokine concentrations with impaired glucose tolerance and type 2 diabetes: results from the Cooperative Health Research in the Region of Augsburg Survey S4 (KORA S4). Diabetes. 2005;54(suppl_2):S11–S7.

35. Dworacka M, Krzyżagórska E, Iskakova S, Bekmukhambetov Y, Urazayev O, Dworacki G. Increased circulating RANTES in type 2 diabetes. European Cytokine Network. 2014;25:46–51.

36. Kaser A, Dunzendorfer S, Offner FA, Ryan T, Schwabegger A, Cruikshank WW, et al. A role for IL-16 in the cross-talk between dendritic cells and T cells. The Journal of Immunology. 1999;163(6):3232–8.

37. McWilliam AS, Napoli S, Marsh AM, Pemper FL, Nelson DJ, Pimm CL, et al. Dendritic cells are recruited into the airway epithelium during the inflammatory response to a broad spectrum of stimuli. The Journal of experimental medicine. 1996;184(6):2429–32.

38. Kaser A, Dunzendorfer S, Offner FA, Ludwiczek O, Enrich B, Koch RO, et al. B lymphocyte-derived IL-16 attracts dendritic cells and Th cells. The Journal of Immunology. 2000;165(5):2474–80.

39. Deng H, Li Z, Liu G, Li X, Chen Y, Zhang Y, et al. Elevated serum interferon γ-inducible protein-10 in women with polycystic ovary syndrome. Gynecological Endocrinology. 2017;33(5):363–7.

40. Herder C, Hauner H, Kempf K, Kolb H, Skurk T. Constitutive and regulated expression and secretion of interferon-γ-inducible protein 10 (IP-10/CXCL10) in human adipocytes. International journal of obesity. 2007;31(3):403–10.

41. Sajadi S, Khoramdelazad H, Hassanshahi G, Rafatpanah H, Hosseini J, Mahmoodi M, et al. Plasma levels of CXCL1 (GRO-alpha) and CXCL10 (IP-10) are elevated in type 2 diabetic patients: evidence for the involvement of inflammation and angiogenesis/angiostasis in this disease state. Clin Lab. 2013;59(1-2):133–7.

42. Seifert HA, Collier LA, Benkovic SA, Willing AE, Pennypacker KR. Abstract WP265: Interferon-inducible Protein 10 Increases following Stroke. Am Heart Assoc; 2013.

43. Coma G, Pena R, Blanco J, Rosell A, Borras F, Este J, et al. Treatment of monocytes with interleukin (IL)-12 plus IL-18 stimulates survival, differentiation and the production of CXC chemokine ligands (CXCL) 8, CXCL9 and CXCL10. Clinical & Experimental Immunology. 2006;145(3):535–44.

44. Zhong B, Guo S, Yang Z, Han L, Du J, Chen J, et al. Roflumilast reduced the IL-18-induced inflammatory response in fibroblast-like synoviocytes (FLS). ACS omega. 2021;6(3):2149–55.

45. Perrier S, Darakhshan F, Hajduch E. IL-1 receptor antagonist in metabolic diseases: Dr Jekyll or Mr Hyde? FEBS letters. 2006;580(27):6289–94.

46. Luotola K, Pietilä A, Alanne M, Lanki T, Loo B-M, Jula A, et al. Genetic variation of the interleukin-1 family and nongenetic factors determining the interleukin-1 receptor antagonist phenotypes. Metabolism. 2010;59(10):1520–7.

47. Luotola K, Piltonen TT, Puurunen J, Tapanainen JS. IL-1 receptor antagonist levels are associated with glucose tolerance in polycystic ovary syndrome. Clinical Endocrinology. 2016;85(3):430–5.

48. Amjadi F, Zandieh Z, Mehdizadeh M, Ajdary M, Aghamajidi A, Raoufi E, et al. Molecular signature of immunological mechanism behind impaired endometrial receptivity in polycystic ovarian syndrome. Archives of Endocrinology and Metabolism. 2022;66:303–11.

49. Bellora F, Castriconi R, Doni A, Cantoni C, Moretta L, Mantovani A, et al. M-CSF induces the expression of a membrane-bound form of IL-18 in a subset of human monocytes differentiating in vitro toward macrophages. European journal of immunology. 2012;42(6):1618–26.

50. Gluszak O, Stopinska-Gluszak U, Glinicki P, Kapuscinska R, Snochowska H, Zgliczynski W, et al. Phenotype and metabolic disorders in polycystic ovary syndrome. International Scholarly Research Notices. 2012;2012.

