## Supplementary material for "Blood Cytokine Profiling Reveals Distinct Inflammatory Signatures in Phenotypes A and B of Polyendocrine Metabolic Ovarian Syndrome (PMOS; formerly PCOS)": Figure S1

**Supplementary Figure Legend**

**Supplementary Figure S1:** Comparative analysis of 34 cytokines that did not show a significant mean difference between control and PMOS groups.

Mean concentration levels (pg/mL) of significantly unaltered cytokines in control and PMOS groups: (A) violin with dot plot representation of unaltered interleukins in control and PMOS groups, (B) representation of unaltered chemokines, (C) representation of unaltered growth factors, and (D) representation of other cytokines in control and PMOS groups. Blue colour represents control subjects and orange colour represents PMOS subjects. Each dot represented a single sample and all 34 cytokines showed p-value > 0.05. Hence, it is considered statistically not significant.
