## Supplementary figures and images for "Blood Cytokine Profiling Reveals Distinct Inflammatory Signatures in Phenotypes A and B of Polyendocrine Metabolic Ovarian Syndrome (PMOS; formerly PCOS)"

### Figure S1

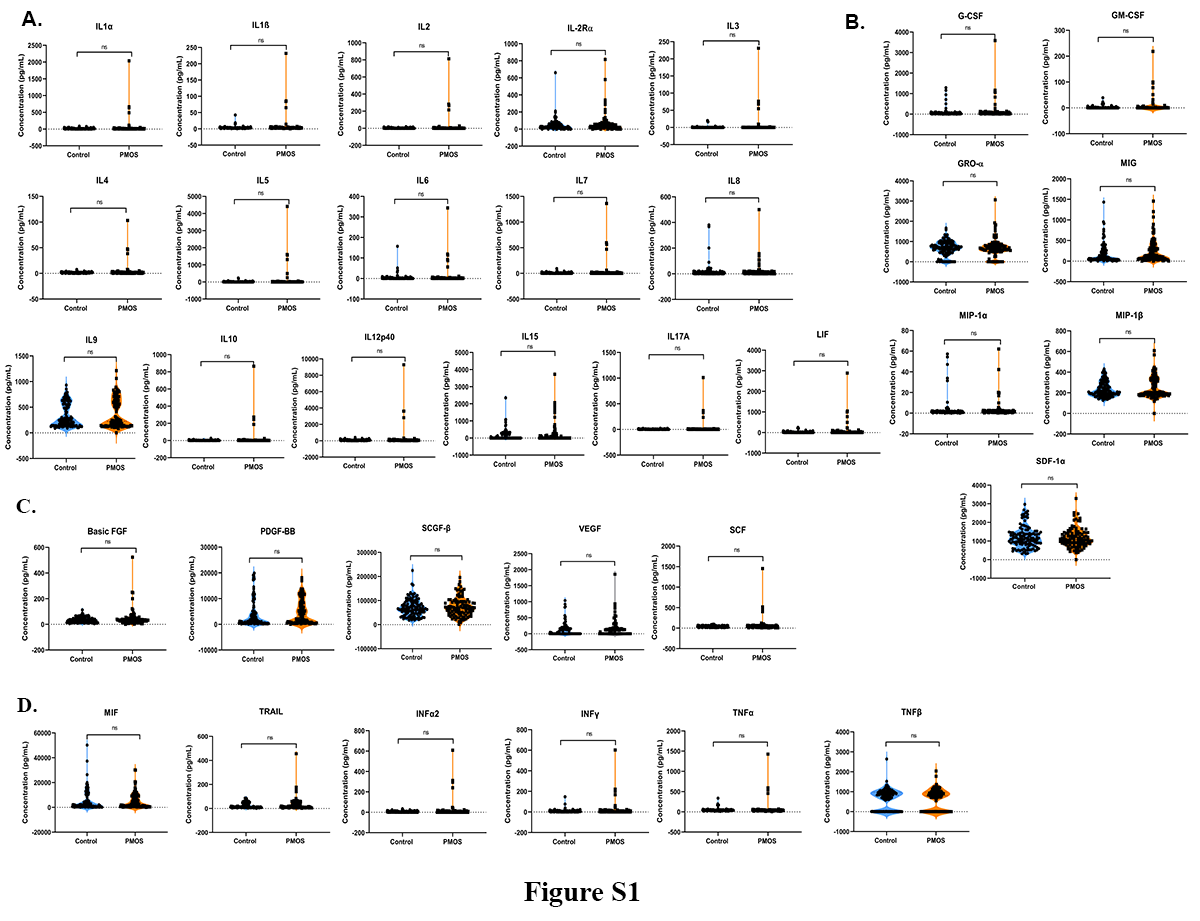

### Figure S2

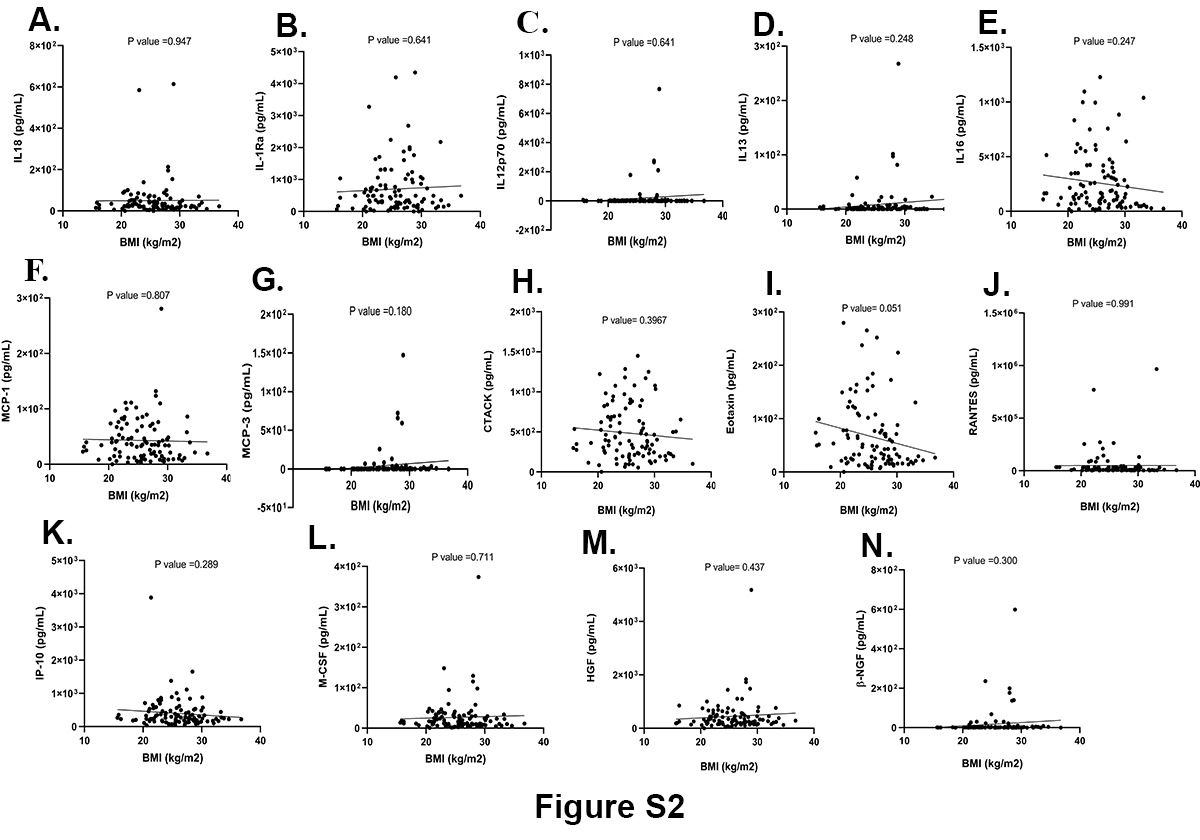
