## Supplementary material for "Blood Cytokine Profiling Reveals Distinct Inflammatory Signatures in Phenotypes A and B of Polyendocrine Metabolic Ovarian Syndrome (PMOS; formerly PCOS)": Figure S2

**Supplementary Figure Legend**

**Supplementary Figure S2:** Linear regression analysis between significantly altered cytokines in the PMOS group with their BMI.

Linear regression analysis of significantly altered cytokines in the PMOS group with their BMI. (A-N) represents the linear regression analysis of IL18, IL-1Ra, IL12p70, IL13, IL16, MCP-1, MCP-3, CTACK, Eotaxin, RANTES, IP-10, M-CSF, HGF and β-NGF respectively. A p-value <0.05 is considered statistically significant.
