## Supplementary material for "Blood Cytokine Profiling Reveals Distinct Inflammatory Signatures in Phenotypes A and B of Polyendocrine Metabolic Ovarian Syndrome (PMOS; formerly PCOS)": Table S1

**Supplementary Table**

**Table S1: List of cytokines that didn’t show a significant mean difference between Control (n=114) and PMOS (n=102) subjects.**

| Cytokines | | Control  (Mean ± SEM) | PMOS  (Mean ± SEM) | p-value  (>0.05) |
| --- | --- | --- | --- | --- |
| Interleukins | IL-1α | 13.1 ± 1.5 | 48.5 ± 22.1 | 0.09 |
|  | IL-1β | 3.7 ± 0.4 | 7.6 ± 2.6 | 0.12 |
|  | IL2 | 2.3 ± 0.4 | 17.2 ± 8.9 | 0.08 |
|  | IL2RA | 47.7 ± 6.5 | 64.8 ± 10.8 | 0.17 |
|  | IL3 | 0.7 ± 0.3 | 4.5 ± 2.5 | 0.11 |
|  | IL4 | 1.6 ± 0.1 | 3.8 ± 1.2 | 0.06 |
|  | IL5 | 10.7 ± 2.9 | 98.8 ± 49.8 | 0.06 |
|  | IL6 | 4.3 ± 1.5 | 9.3 ± 3.9 | 0.21 |
|  | IL7 | 6.4 ± 1.2 | 34.1 ± 15.9 | 0.07 |
|  | IL8 | 17.4 ± 5 | 19.1 ± 5.4 | 0.83 |
|  | IL9 | 333.4 ± 20.9 | 363.3 ± 27.2 | 0.38 |
|  | IL10 | 1.9 ± 0.3 | 17.1 ± 9.3 | 0.08 |
|  | IL12p40 | 70.1 ± 6.6 | 252.9 ± 105.4 | 0.07 |
|  | IL15 | 138.8 ± 28.3 | 208.7 ± 53.9 | 0.24 |
|  | IL17A | 4.8 ± 0.5 | 23.1 ± 11.1 | 0.08 |
|  | LIF | 21.9 ± 4.2 | 77.8 ± 32.4 | 0.07 |
| Growth Factors | Basic FGF | 33.9 ± 1.9 | 44.7 ± 6.2 | 0.08 |
|  | PDGF BB | 4126.2 ± 498.6 | 4717.1 ± 459.6 | 0.39 |
|  | SCGF-β | 69879.8 ± 3332.5 | 73625.6 ± 3898.1 | 0.46 |
|  | VEGF | 122.9 ± 18.3 | 153.3 ± 25.6 | 0.33 |
|  | SCF | 40.9 ± 2.1 | 68 ± 15.6 | 0.07 |
| Chemokines | G-CSF | 83.3 ± 18.3 | 127.8 ± 39.7 | 0.29 |
|  | GM-CSF | 1.9 ± 0.5 | 6.4 ± 2.7 | 0.88 |
|  | GRO-α | 680.6 ± 33.9 | 696.2 ± 45.7 | 0.78 |
|  | MIG | 178.9 ± 21.7 | 239 ± 26.7 | 0.08 |
|  | MIP-1α | 3.4 ± 0.9 | 3.4 ± 0.8 | 0.95 |
|  | MIP-1β | 233.9 ± 6.8 | 248.1 ± 9.9 | 0.23 |
|  | SDF-1α | 1204 ± 53.3 | 1166.9 ± 51.8 | 0.62 |
| Other Cytokines | MIF | 5394.1 ± 729.9 | 5235.9 ± 604 | 0.87 |
|  | TRAIL | 23.5 ± 2 | 32 ± 5.1 | 0.11 |
|  | INF-α2 | 5.3 ± 0.7 | 19.3 ± 7.5 | 0.05 |
|  | INF-γ | 8.3 ± 1.5 | 17.8 ± 6.7 | 0.15 |
|  | TNF-α | 46.8 ± 3.8 | 65.6 ± 15.9 | 0.23 |
|  | TNF-β | 629.2 ± 46.4 | 607.5 ± 50.1 | 0.75 |

List of 34 cytokines that didn’t show a significant mean difference between Control (n=114) and PMOS (n=102) subjects. All values were presented as Mean ± SEM. The concentration of cytokines is expressed in pg/mL. A p-value > 0.05 is considered as statistically insignificant.
