## Supplementary material for "Blood Cytokine Profiling Reveals Distinct Inflammatory Signatures in Phenotypes A and B of Polyendocrine Metabolic Ovarian Syndrome (PMOS; formerly PCOS)": Table S2

**Supplementary Table**

**Table S2: Clinical characteristics and cytokine levels in individuals with four different PMOS phenotypes.**

| Characteristics/ Cytokines | Phenotype A (52.9%) | Phenotype B (4.9%) | Phenotype C (31.4%) | Phenotype D (10.8%) |
| --- | --- | --- | --- | --- |
| Age (years) | 27.9 ± 0.5 | 25 ± 1.1 | 27.9 ± 0.8 | 27 ± 1.3 |
| BMI (kg/m^2^) | 25.6 ± 0.5 | 21 ± 2.4 | 26 ± 0.8 | 27 ± 1.4 |
| LH | 14.4 ± 0.9 | 10.3 ± 2.9 | 17.2 ± 1.4 | 14.6 ± 2.1 |
| FSH | 5.6 ± 0.3 | 5.9 ± 0.7 | 6.8 ± 0.9 | 5.4 ± 0.8 |
| LH/FSH | 3.1 ± 0.2 | 1.8 ± 0.6 | 3.3 ± 0.3 | 3.2 ± 0.6 |
| Testosterone | 48.6 ± 1.7 | 51.6 ± 7 | 55.2 ± 3.3 | 11.4 ± 1.9 |
| DHT | 0.17 ± 0.02 | 0.15 ± 0.06 | 0.17 ± 0.02 | 0.15 ± 0.04 |
| Ovary size (cm^3^) | 11.6 ± 0.7 | 9.4 ± 1.9 | 10.9 ± 0.7 | 10.1± 1.3 |
| IL18 (pg/mL) | 51.4 ± 10.8 | 144.6 ± 117.4 | 41.9 ± 9.4 | 24.7 ± 4.7 |
| IL-1Ra (pg/mL) | 751.2 ± 114.3 | 1260.5 ± 791.6 | 522.7 ± 100.9 | 726.9 ± 178.1 |
| IL12p70 (pg/mL) | 6.4 ± 3.4 | 154.2 ± 153.3 | 25.4 ± 13.1 | 1 ± 0.4 |
| IL13 (pg/mL) | 4.4 ± 1.3 | 54.7 ± 53.2 | 11.2 ± 4.8 | 2.7 ± 2 |
| IL16 (pg/mL) | 282.9 ± 40.9 | 356.5 ± 152.7 | 192.1 ± 37.5 | 280.3 ± 80.7 |
| MCP-1(CCL2) (pg/mL) | 42.9 ± 4.1 | 81.2 ± 50.3 | 38.9 ± 6.5 | 37.8 ± 8.1 |
| MCP-3 (CCL7) (pg/mL) | 1.2 ± 0.5 | 30.9 ± 29.1 | 6.7 ± 3.4 | 1.2 ± 1.1 |
| CTACK(CCL27) (pg/mL) | 570.9 ± 50.5 | 629.7 ± 169.9 | 351.2 ± 43.5 | 432.4 ± 87.5 |
| Eotaxin (CCL11) (pg/mL) | 70.3 ± 8.4 | 74.6 ± 28.1 | 58.7 ± 11.8 | 58.1 ± 19.2 |
| RANTES(CCL5) (pg/mL) | 65232.5 ± 24545.7 | 24479.1 ± 6382.9 | 37299.7 ± 12091.6 | 26117.1 ± 11177.1 |
| IP-10(CXCL-10) (pg/mL) | 471.6 ± 78.7 | 339.5 ± 55.9 | 304.4 ± 40.9 | 367.2 ± 69.4 |
| M-CSF(CSF-1) (pg/mL) | 25.3 ± 3.3 | 87.7 ± 71.6 | 24.1 ± 5.8 | 15.9 ± 2.9 |
| HGF (pg/mL) | 430.2 ± 38.7 | 1223.4 ± 990.9 | 407.4 ± 82.6 | 323.1 ± 68.2 |
| β-NGF (pg/mL) | 8.8 ± 4.5 | 127.1 ± 118.1 | 23.4 ± 9.7 | 3.2 ± 2.7 |

Plasma level of physiological characters, hormones, and significantly altered cytokines in four PMOS phenotype sub-groups. All values were presented as Mean ± SEM. Each PMOS phenotype is represented by the percentage of the population in this study group.
