## Supplementary material for "Blood Cytokine Profiling Reveals Distinct Inflammatory Signatures in Phenotypes A and B of Polyendocrine Metabolic Ovarian Syndrome (PMOS; formerly PCOS)": Table S3

**Supplementary Table**

**Table S3: Odds ratio calculation of altered cytokines in control (n=114) and PMOS (n=102) subjects.**

| Cytokines | Lower 95% CI | Odds Ratio (OR) | Upper 95% CI |
| --- | --- | --- | --- |
| IL18 | 1.11 | 1.92 | 3.33 |
| IL-1Ra | 1.15 | 2 | 3.47 |
| IL12p70 | 0.39 | 0.7 | 1.25 |
| IL13 | 0.71 | 1.24 | 2.16 |
| IL16 | 1.65 | 2.91 | 5.15 |
| MCP-1(CCL2) | 1.33 | 2.29 | 3.96 |
| MCP-3(CCL7) | 0.37 | 0.69 | 1.27 |
| CTACK(CCL27) | 0.87 | 1.49 | 2.56 |
| Eotaxin(CCL11) | 0.85 | 1.47 | 2.52 |
| RANTES | 0.76 | 1.31 | 2.25 |
| IP-10 (CXCL10) | 0.89 | 1.53 | 2.63 |
| M-CSF (CSF-1) | 1.15 | 2.01 | 3.49 |
| HGF | 0.95 | 1.65 | 2.85 |
| Β-NGF | 0.39 | 0.78 | 1.51 |

Odds ratio calculation along with a lower 95% class interval (CI), and upper 95% CI of 14 significantly altered cytokines in control (n=114) and PMOS (n=102) subjects.
