## Supplementary material for "Blood Cytokine Profiling Reveals Distinct Inflammatory Signatures in Phenotypes A and B of Polyendocrine Metabolic Ovarian Syndrome (PMOS; formerly PCOS)": Table S4

**Supplementary Table**

**Table S4: Top 10 significant pairwise correlation cytokine combinations in the PMOS group.**

| Sl No | Cytokine1 | Cytokine 2 | Correlation coefficient (R) | p-value |
| --- | --- | --- | --- | --- |
| 1 | IL16 | IL-1Ra | 0.75 | < 0.0001 |
| 2 | IL18 | IL13 | 0.70 | < 0.0001 |
| 3 | IL18 | M-CSF | 0.91 | < 0.0001 |
| 4 | IL18 | HGF | 0.77 | < 0.0001 |
| 5 | IL13 | MCP-1 | 0.71 | < 0.0001 |
| 6 | IL13 | M-CSF | 0.86 | < 0.0001 |
| 7 | IL13 | HGF | 0.89 | < 0.0001 |
| 8 | MCP-1 | M-CSF | 0.76 | < 0.0001 |
| 9 | MCP-1 | HGF | 0.79 | < 0.0001 |
| 10 | M-CSF | HGF | 0.91 | < 0.0001 |

Top 10 significant Pearson’s pairwise correlation cytokine combinations in the PMOS group. All cytokine combinations are represented as correlation coefficient (R) ≥ 0.7 and p-value <0.05.
